# Biliary Duct Hamartomas: A Systematic Review

**DOI:** 10.1101/2021.04.27.21256209

**Authors:** Abu Baker Sheikh, Rahul Shekhar, Nismat Javed, Katarina Leyba, Abdul Ahad Ehsan Sheikh, Shubhra Upadhyay, Devika Kapuria, Christopher Cormier, Tarun Rustagi

**Affiliations:** University of New Mexico Health Sciences Center, Department of Internal Medicine, Albuquerque, NM, USA..; Division of Hospital Medicine, University of New Mexico Health Sciences Center, Department of Internal Medicine, Albuquerque, NM, USA..; Shifa College of Medicine, Shifa Tameer-e-Millat University, Islamabad, Pakistan..; University of New Mexico, Department of Internal Medicine, Albuquerque, NM, USA..; The Wright Center for Graduate Medical Education, Department of Internal Medicine, Scranton, PA, USA..; Clinical Fellow Department of Gastroenterology and Hepatology, University of New Mexico Health Science Center, Albuquerque, NM, USA..; Resident Physician Department of Pathology, University of New Mexico Health Sciences Center, Albuquerque, New Mexico, USA..; Interventional Endoscopist & Assistant Professor Division of Gastroenterology and Hepatology, University of New Mexico Health Sciences Center, Albuquerque, New Mexico, USA..

**Keywords:** Biliary duct hamartomas, clinical presentation, VMC, imaging

## Abstract

Biliary duct hamartomas are benign lesions of intrahepatic bile ducts. Oftentimes an incidental finding on imaging, these lesions can pose a diagnostic dilemma because of their overlapping features with malignant masses. We performed a structured systematic review of literature and identified 139 cases of biliary duct hamartomas. Patient demographics, clinical presentation with key laboratory and imaging findings, diagnostic modalities, management strategies, and outcomes were analyzed and systematically summarized. The systematic review is aimed to help with better understanding of biliary duct hamartomas and its principal features.

## INTRODUCTION

Biliary duct hamartomas, or “von Meyenberg complexes,” are benign malformations of intrahepatic bile ducts.^1, 2^ They are often found incidentally on imaging or laparotomy, and although clinically benign, they can mimic malignant lesions, and present with clinical symptoms that prompts a lengthy diagnostic workup and even surgical intervention. The rare nature of this condition as well as heterogenous clinical presentation can make management challenging^1, 3^, and it is therefore important to recognize biliary hamartomas and distinguish them from other pathologies.

Overall, biliary hamartomas are relatively rare findings that result from the failure of involution of embryonic bile ducts, typically malformations of smaller interlobular bile ducts. Their estimated prevalence ranges from 0.6-5.6% on autopsy and is around 1.0% on imaging.^4, 5^ Although biliary duct hamartomas have been described in children, they most commonly affect individuals age 35 and older.^1^ They are more common in patients with polycystic liver disease and polycystic kidney disease,^6^ and studies have found that they occur at increased frequency in patients with cirrhosis, indicating that there may be some component of environmental exposure involved in their pathogenesis.^1, 7^

Most biliary hamartomas are asymptomatic, although they can uncommonly cause fever, jaundice or abdominal pain. Similarly, most patients present with an unremarkable clinical exam and normal lab results, but rare elevations in transaminases, alkaline phosphatase or gamma-glutamyl transferase (GGT) levels can occur.^1^ While their clinical course is generally benign, as with other ductal plate malformations, biliary hamartomas do pose a small risk of malignant transformation to intrahepatic cholangiocarcinoma or, less frequently, hepatocellular carcinoma.^8^ Given the marked differences between the management of biliary hamartomas and other, similar-appearing diseases, it is important to fully characterize them in order to facilitate accurate diagnosis and optimal management. Within this context, our systematic review aims to provide a comprehensive clinical profile of biliary duct hamartomas and an up to date overview of all cases reported in the literature to identify salient and distinguishing features.

## METHODS

### Protocol development and systematic review registration

We developed the protocol for this review after consensus with all the reviewers and subject experts per PRISMA guidelines. After protocol preparation and registration on PROSPERO (CRD42021230745), data search was performed as below.

### Search strategy

Keywords (including all commonly used abbreviations of these terms) used in the search strategy were as follows: “Biliary hamartomas” OR “Von Meyenburg Complexes” OR “Biliary duct hamartomas” OR “Hamartomas AND biliary” OR “Multiple biliary hamartomas” OR “Multiple biliary duct hamartomas”.

### Data extraction (selection and coding)

We screened PubMed, Web of Science and CINAHL databases and articles published in the English language were included in the systemic review over the last 30 years. On the initial search, 163 articles on PubMed, 33 on Web of Science (excluding Medline) and 63 on CINHAL (excluding Medline) were included. After the initial search, duplicates were removed, and we imported all included search study into EndNote online software (Figure 1). Two independent reviewers (NJ and ABS) screened remaining studies for the inclusion based on inclusion criteria, and researchers were blinded to each other’s decisions. Rayyan software and Mendeley desktop were used. The screening was done via reading the abstract and if needed by reading full-text articles. Studies published in the English language or English translation as well as available for viewing were included in the initial review. Once the initial screening was done, two independent reviewers (SU and AAES) reviewed the full-text article for final inclusion. Reviewers were blinded to each other’s decisions, and a third reviewer resolved any dispute (RS).

**Figure 1:**
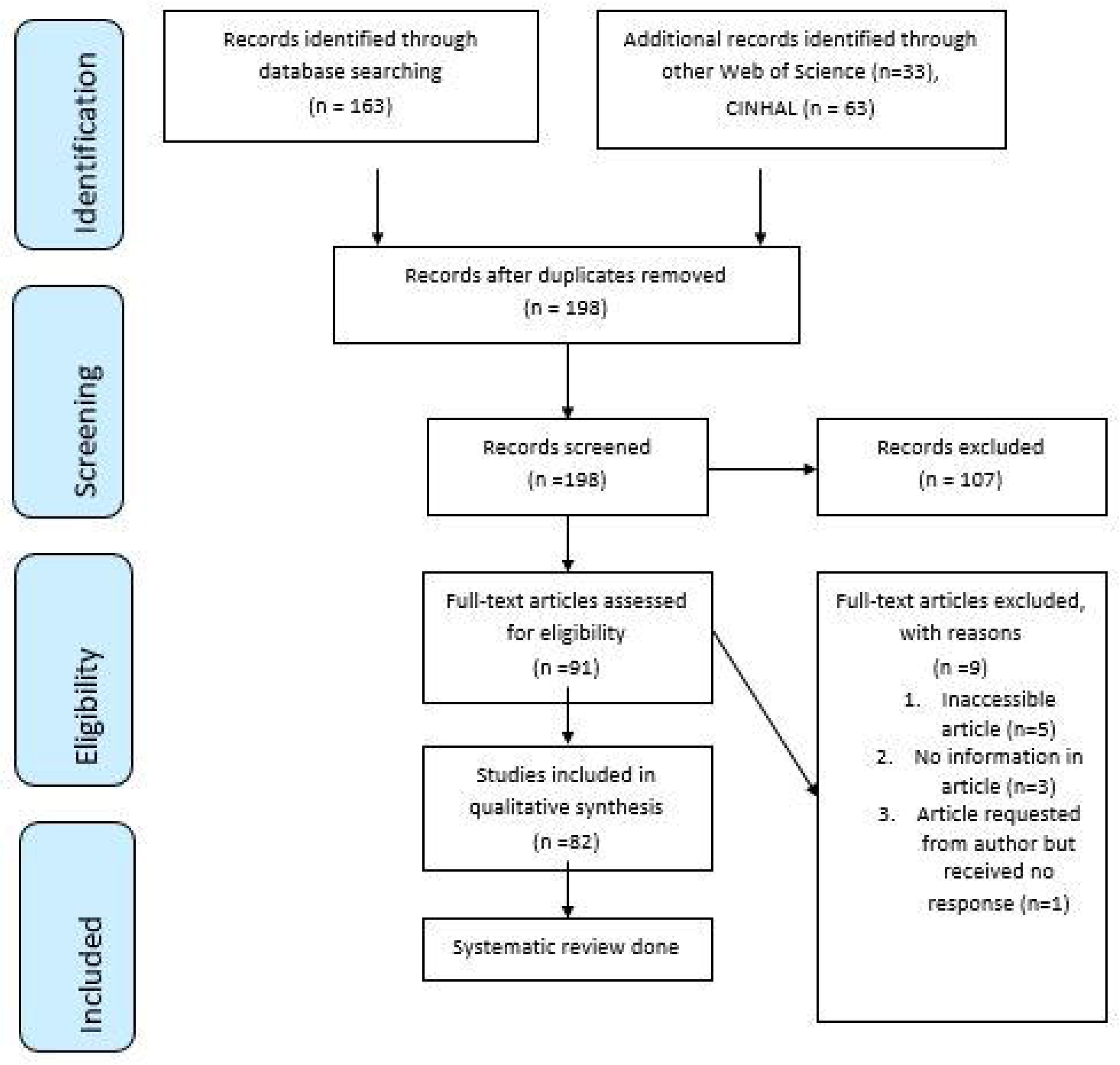
PRISMA flow chart.

Data was extracted from study documents, including information about study design and methodology, participant demographics and baseline characteristics, study country, publication journal, clinical presentation, symptoms, laboratory data, imaging data, intervention, treatment, clinical outcomes, morbidity, and mortality.

One reviewer did data extraction, and another reviewer cross-checked the extracted data for accuracy and completeness. Any disagreements between individual judgements were resolved via the third reviewer. Attempts were made to obtain any missing data from study corresponding investigators via email. If data could not be obtained, that study was excluded from the analysis on a case-by-case basis. Publications that were not peer-reviewed were excluded from this study. The preliminary data was entered and recorded in an excel spreadsheet. The final analysis included 82 studies.

### Risk of bias (quality) assessment

Quality assessment of all the included studies was assessed using the methodological quality and synthesis of case series and case reports described by Murad et al. (2018).^9^ (Table 1 and Table 2)

According to this tool, four broad perspectives to assess the quality; selection of the study groups, ascertainment of the observed outcome, causality of the observed outcome and case reporting

### Strategy for data synthesis

For statistical analysis, we used IBM SPSS Statistics version 21 (IBM, Armonk, NY, USA). Based on the distribution of values, continuous data were expressed as mean ± standard deviation. Qualitative variables were expressed as frequency and percentages.

## RESULTS

To date, 139 patients with bile duct hamartomas have been reported in the literature, including 68 case reports and 15 case series. These 139 patients were included in the analysis. Summarized data from all analyzed cases are reported in Tables 3, 4 and 5.^2, 7, 8, 10–89^

**Table 3:**
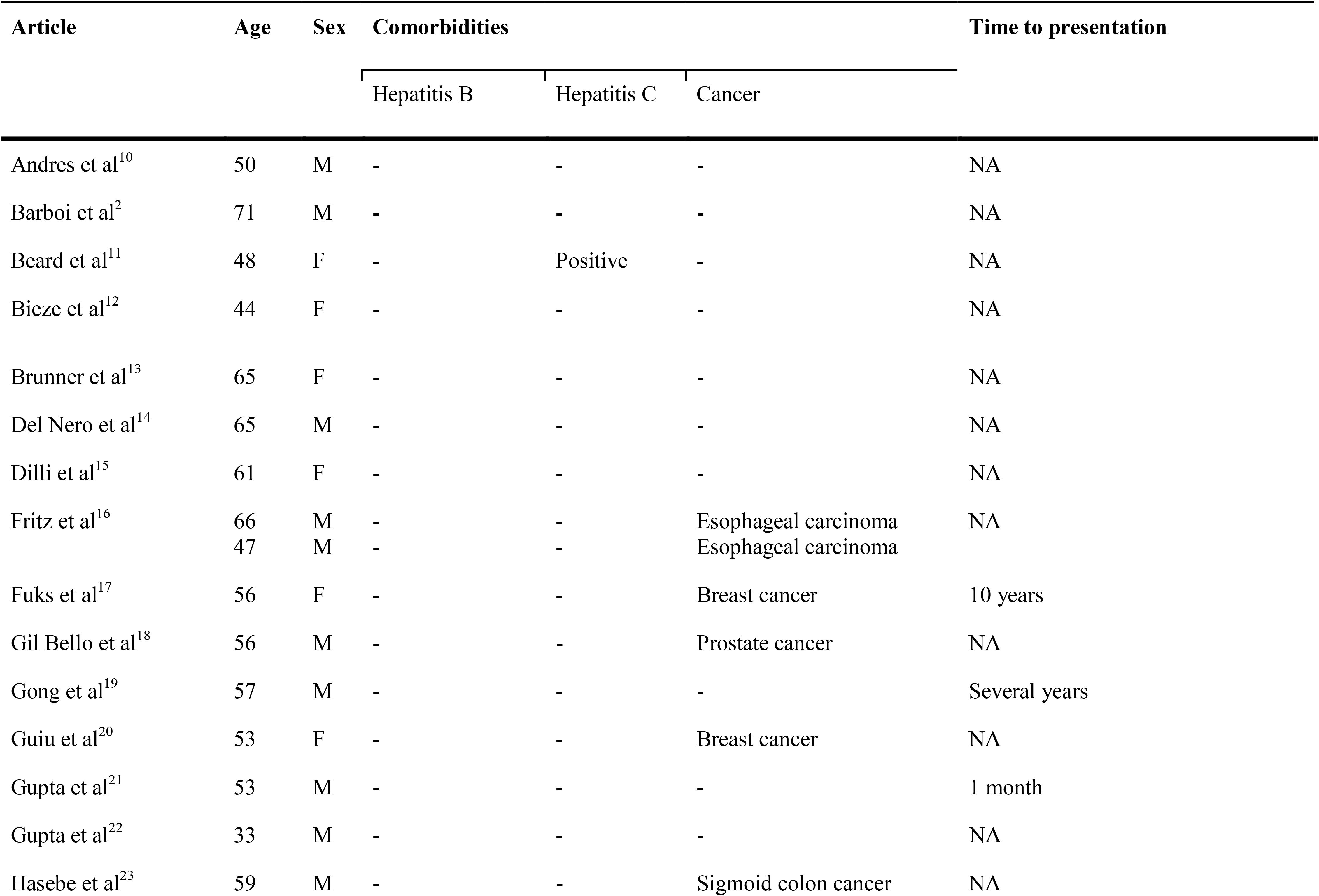
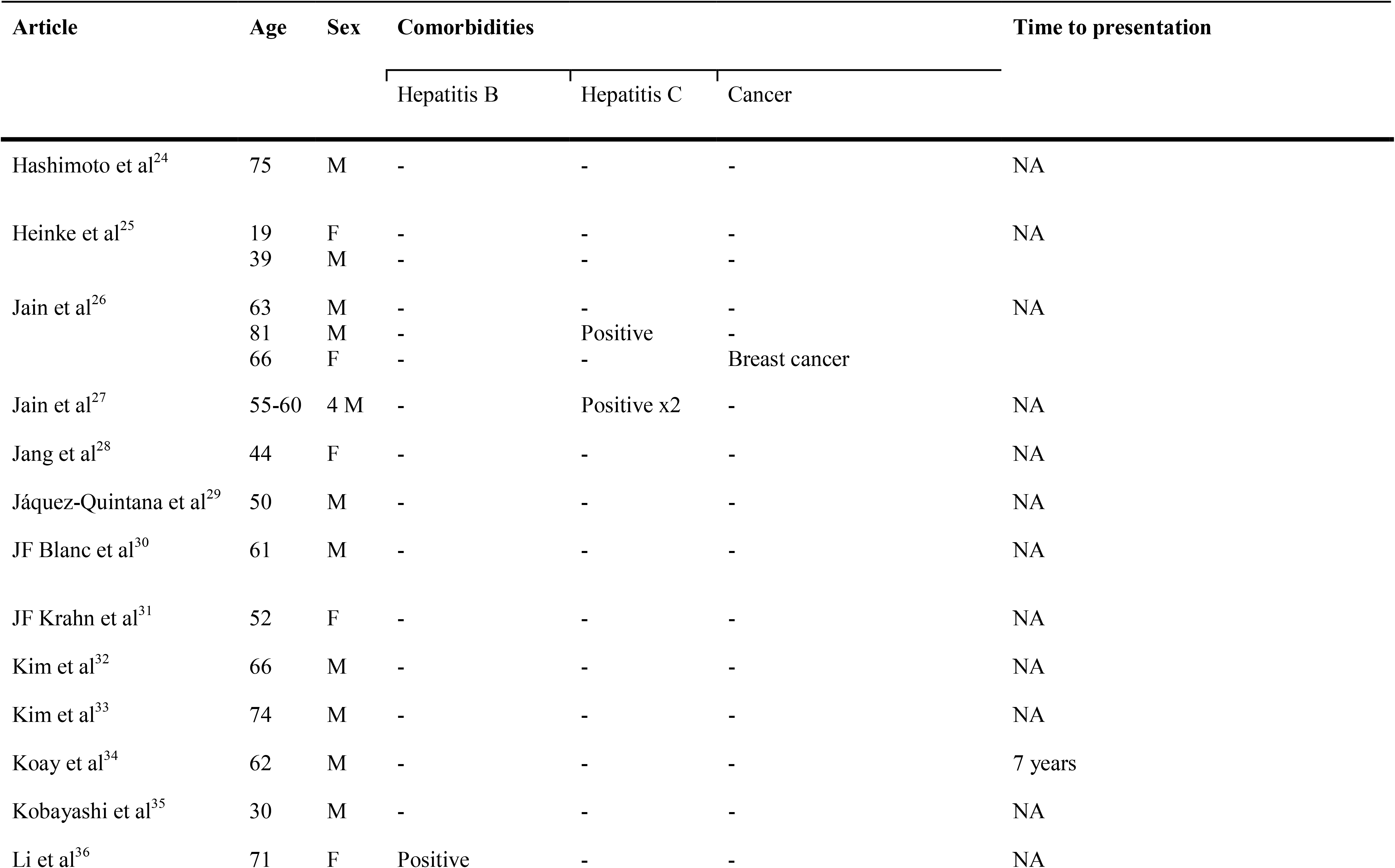
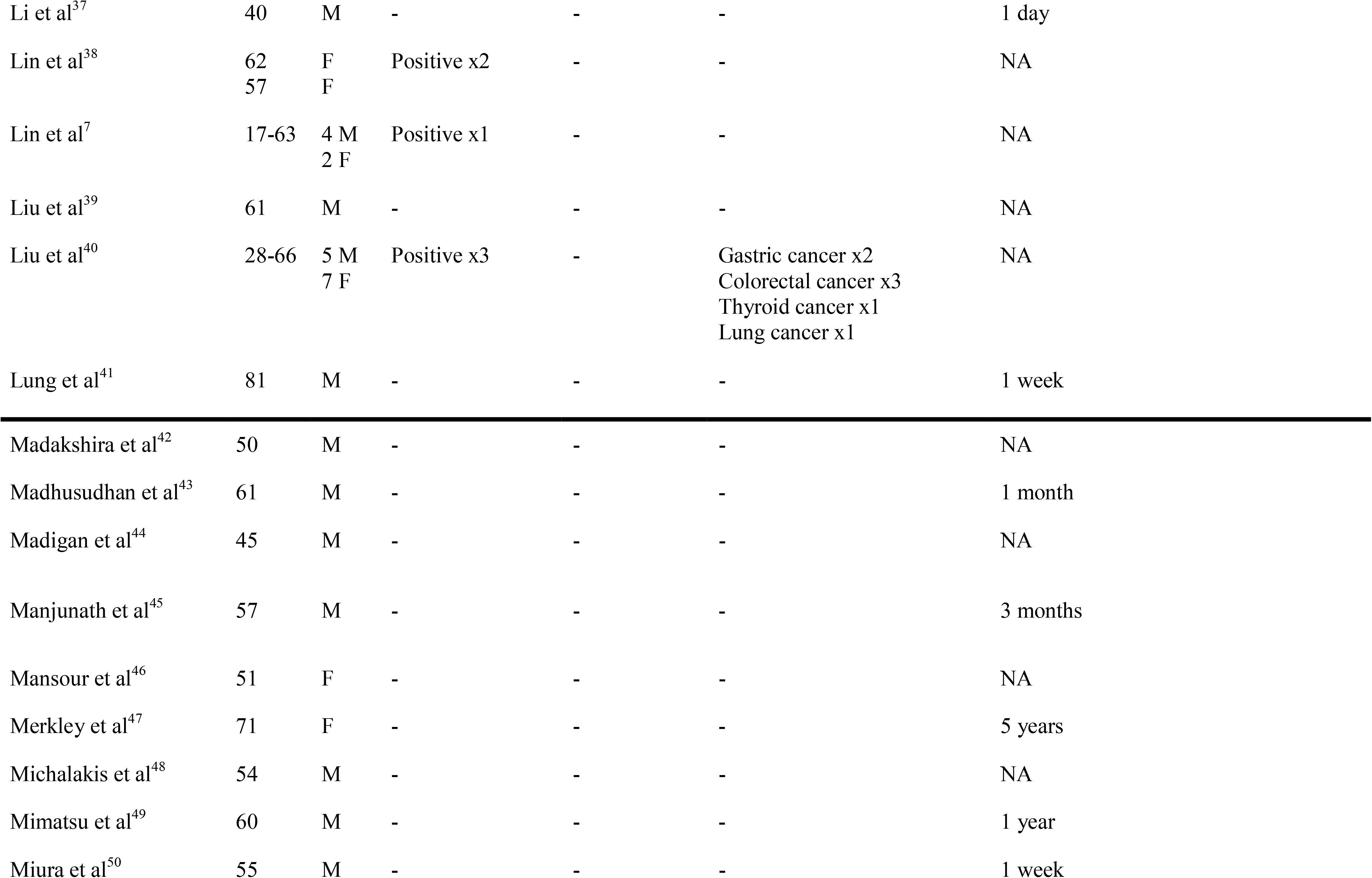
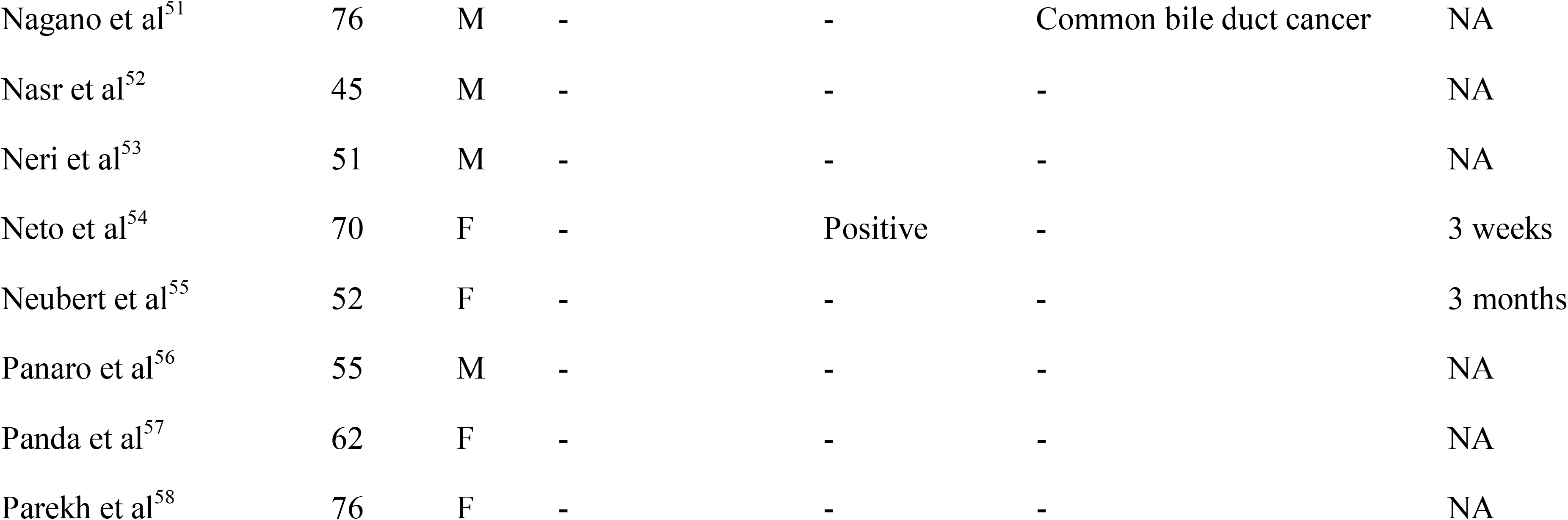
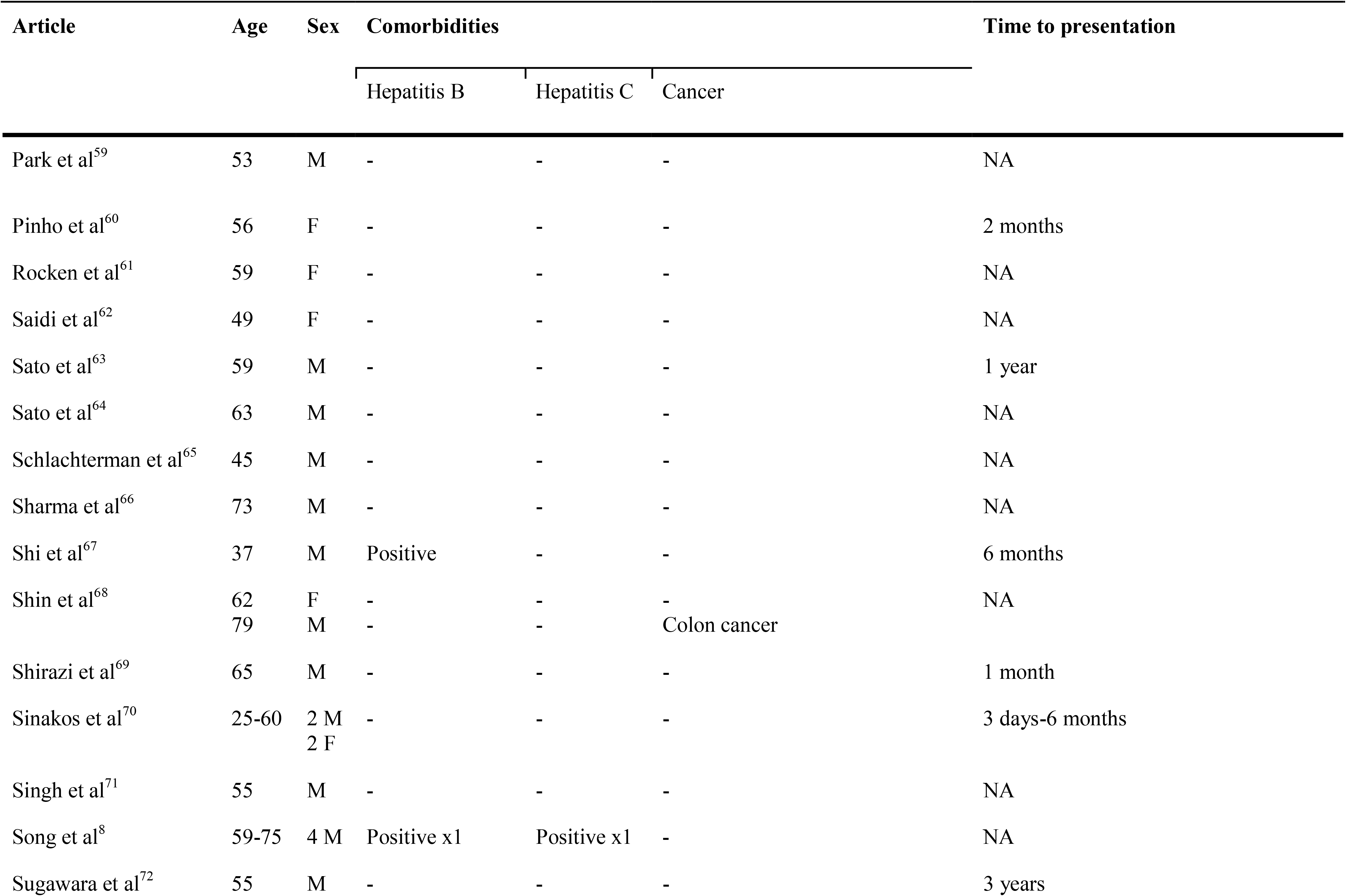
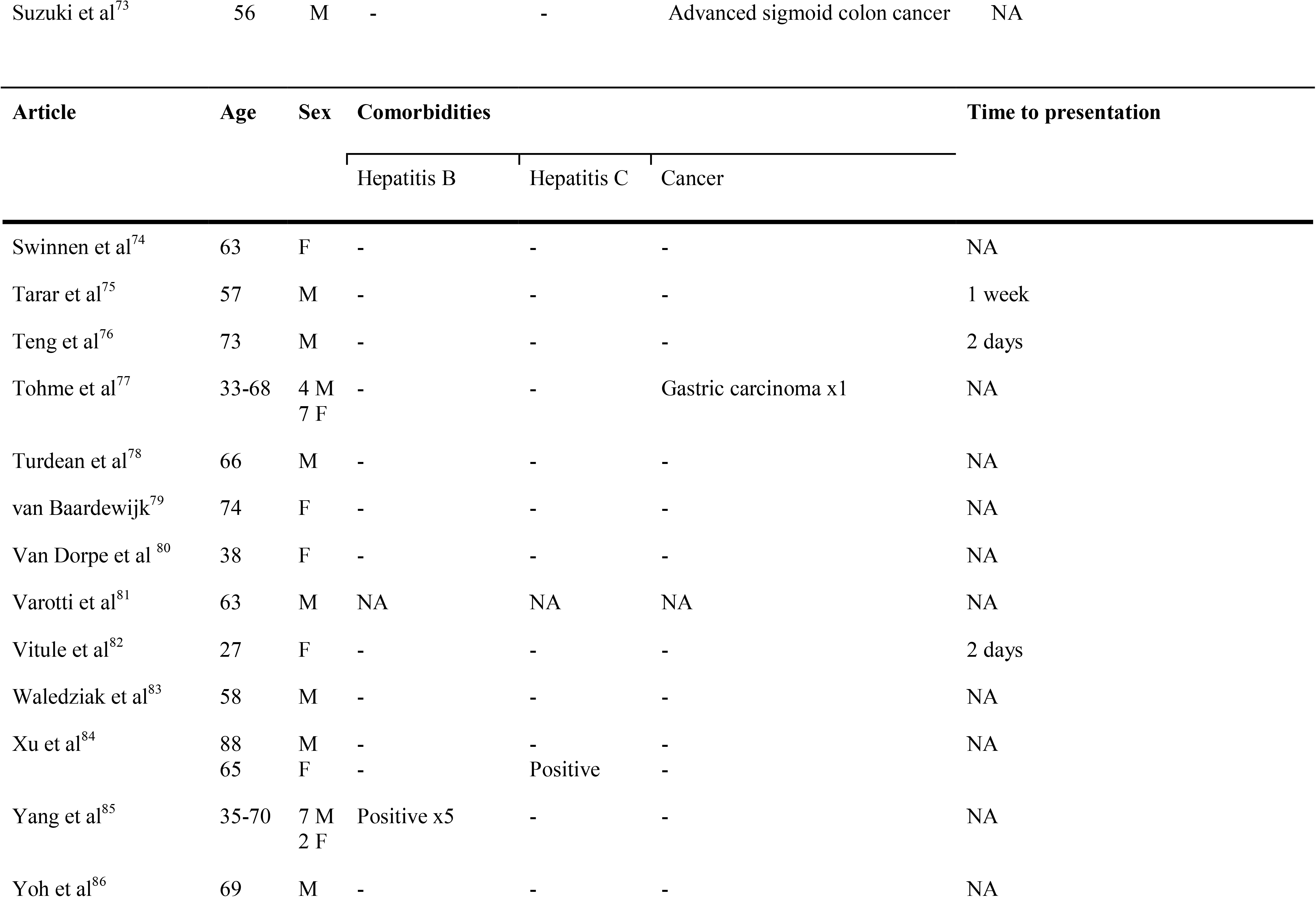
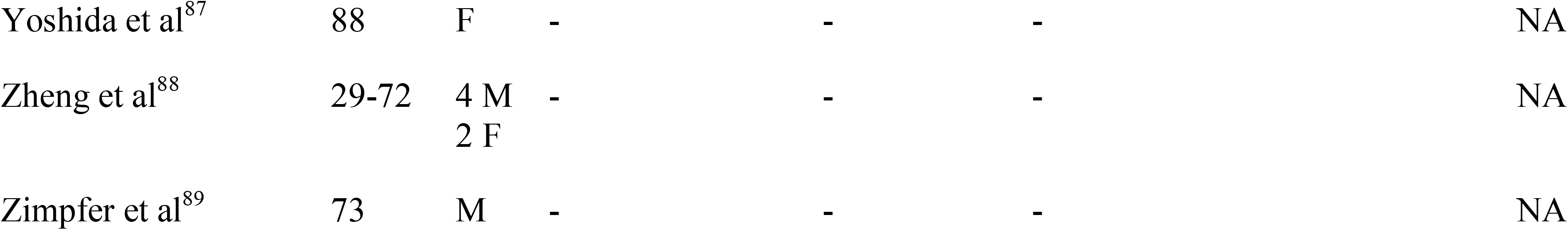
Demographic characteristics of the cases reviewed.

**Table 4:** Clinical presentation and laboratory findings of the cases reviewed.

| Article | Symptoms |  |  | Labs |  |  | Clinical exam |
| --- | --- | --- | --- | --- | --- | --- | --- |
|  | Abdominal pain | Jaundice | Fever | AST/ALT | GGT | Bilirubin |  |
| Andres et al<sup>10</sup> | - | - | - | Elevated | Elevated | - | NA |
| Barboi et al<sup>2</sup> | - | - | - | - | - | - | Hepatomegaly |
| Beard et al<sup>11</sup> | Present | - | - | - | - | - | NA |
| Bieze et al<sup>12</sup> | Present | - | - | Elevated | - | - | NA |
| Brunner et al<sup>13</sup> | - | - | - | - | - | - | NA |
| Del Nero et al<sup>14</sup> | - | - | - | - | Elevated | - | Hepatomegaly |
| Dilli et al<sup>15</sup> | Present | - | - | - | - | - | NA |
| Fritz et al<sup>16</sup> | - | - | - | NA | NA | NA | NA |
| Fuks et al<sup>17</sup> | - | - | - | NA | NA | NA | NA |
| Gil Bello et al<sup>18</sup> | - | - | - | - | - | - | NA |
| Gong et al<sup>19</sup> | Present | - | - | Elevated ALT only | - | Elevated | NA |
| Guiu et al<sup>20</sup> | - | - | - | - | - | - | NA |
| Gupta et al<sup>21</sup> | - | Present | - | Elevated | Elevated | Elevated | Jaundice, hepatomegaly |
| Gupta et al<sup>22</sup> | Present | - | - | - | - | - | NA |
| Hasebe et al<sup>23</sup> | - | - | - | - | - | - | NA |
| Hashimoto et al<sup>24</sup> | - | - | Present | Elevated | Elevated | - | NA |
| Heinke et al<sup>25</sup> | - | Present | - | - | - | - | NA |
| Jain et al<sup>26</sup> | Present x2 | - | - | Elevated<br>ALT only x1 | - | - | Splenomegaly x2, ascites x2,<br>varices x1, portal HTN x1 |
|  | Abdominal pain | Jaundice | Fever | AST/ALT | GGT | Bilirubin |  |
| Jain et al<sup>27</sup> | Present x2 | - | Present x1 | - | - | - | NA |
| Jang et al<sup>28</sup> | - | - | - | - | - | Elevated | NA |
| Jáquez-Quintana et al<sup>29</sup> | - | - | - | Elevated<br>AST only | Elevated | - | NA |
| JF Blanc et al<sup>30</sup> | - | - | - | - | Elevated | - | NA |
| JF Krahn et al<sup>31</sup> | - | - | - | - | Elevated | - | HTN |
| Kim et al<sup>32</sup> | - | - | - | - | Elevated | - | NA |
| Kim et al<sup>33</sup> | - | - | - | - | - | - | NA |
| Koay et al<sup>34</sup> | Present | Present | Present | NA | NA | NA | Positive Murphy's sign, tenderness in RUQ |
| Kobayashi et al<sup>35</sup> | - | - | - | NA | NA | NA | NA |
| Li et al<sup>36</sup> | - | - | - | - | - | - | NA |
| Li et al<sup>37</sup> | - | - | - | - | - | - | NA |
| Lin et al<sup>38</sup> | - | - | - | Elevated x2 | - | - | NA |
| Lin et al<sup>7</sup> | - | Present x1 | - | Elevated x1 | - | - | NA |
| Liu et al<sup>39</sup> | - | - | - | - | - | - | NA |
| Liu et al<sup>40</sup> | - | - | - | - | - | - | NA |
| Lung et al<sup>41</sup> | Present | - | - | - | - | - | Abdominal distension and<br>guarding |
|  | Abdominal pain | Jaundice | Fever | AST/ALT | GGT | Bilirubin |  |
| Madakshira et al<sup>42</sup> | - | - | - | - | - | - | NA |
| Madhusudhan et al<sup>43</sup> | - | - | - | - | - | - | NA |
| Madigan et al<sup>44</sup> | - | - | - | - | - | - | Splenomegaly, gynecomastia, +1 pedal edema |
| Manjunath et al<sup>45</sup> | Present | - | - | - | - | - | Hepatomegaly |
| Mansour et al<sup>46</sup> | - | - | - | Elevated ALT only | Elevated | - | NA |
| Merkley et al<sup>47</sup> | Present | - | - | - | - | - | NA |
| Michalakis et al<sup>48</sup> | - | - | - | - | - | - | NA |
| Mimatsu et al<sup>49</sup> | - | - | - | - | - | - | NA |
| Miura et al<sup>50</sup> | Present | Present | - | Elevated | Elevated | Elevated | Jaundice |
| Nagano et al<sup>51</sup> | Present | - | - | - | - | - | NA |
| Nasr et al<sup>52</sup> | Present x1 | - | - | - | - | - | Positive Murphy's sign x1, RUQ pain x1 |
| Neri et al<sup>53</sup> | Present | - | - | - | - | - | Hepatomegaly |
| Neto et al<sup>54</sup> | Present | - | - | Elevated | - | - | Positive Murphy's sign, RUQ tenderness |
| Neubert et al<sup>55</sup> | - | Present | Present | - | - | Elevated | HoTN, fever, tachycardia |
| Panaro et al<sup>56</sup> | - | - | - | - | - | - | NA |
| Panda et al<sup>57</sup> | - | - | - | - | - | - | NA |
| Parekh et al<sup>58</sup> | - | - | - | - | - | - | NA |
|  | Abdominal pain | Jaundice | Fever | AST/ALT | GGT | Bilirubin |  |
| Park et al<sup>59</sup> | - | - | - | - | - | - | NA |
| Pinho et al<sup>60</sup> | - | - | - | - | - | - | NA |
| Rocken et al<sup>61</sup> | Present | - | - | - | Elevated | - | NA |
| Saidi et al<sup>62</sup> | - | - | - | Elevated | - | - | NA |
| Sato et al<sup>63</sup> | - | - | Present | Elevated | - | - | Bilateral peripheral edema and distal polyneuropathy |
| Sato et al<sup>64</sup> | Present | - | - | - | - | - | NA |
| Schlachterman et al<sup>65</sup> | - | - | - | Elevated ALT only | Elevated | - | NA |
| Sharma et al<sup>66</sup> | - | - | - | - | - | - | NA |
| Shi et al<sup>67</sup> | - | - | - | NA | NA | NA | NA |
| Shin et al<sup>68</sup> | - | - | - | - | - | - | Weight loss |
| Shirazi et al<sup>69</sup> | Present | - | - | - | - | Elevated | Vague mass in the RUQ |
| Sinakos et al<sup>70</sup> | Present x4 | Present x1 | Present x2 | Elevated x2 | Elevated x1 | - | RUQ tenderness x2 |
| Singh et al<sup>71</sup> | Present | - | - | - | - | - | Vague mass in the LUQ |
| Song et al<sup>8</sup> | - | - | - | NA | NA | NA | NA |
| Sugawara et | - | - | Present | - | - | - | NA |

al<sup>72</sup> Suzuki et al<sup>73</sup> - - - - - - NA
| Article | Symptoms |  |  | Labs |  |  | Clinical exam |
| --- | --- | --- | --- | --- | --- | --- | --- |
|  | Abdominal pain | Jaundice | Fever | AST/ALT | GGT | Bilirubin |  |
| Swinnen et al<sup>74</sup> | Present | - | - | NA | NA | NA | Normal physical exam |
| Tarar et al<sup>75</sup> | - | - | Present | - | - | - | NA |
| Teng et al<sup>76</sup> | Present | - | Present | - | - | - | Positive Murphy's sign, RUQ tenderness |
| Tohme et al<sup>77</sup> | - | - | - | - | Elevated | - | NA |
| Turdean et al<sup>78</sup> | Present | - | - | - | - | - | NA |
| van Baardewijk<sup>79</sup> | - | - | - | - | - | - | Friable tumor on rectal exam |
| Van Dorpe et al<sup>80</sup> | Present | - | - | - | - | - | NA |
| Varotti et al<sup>81</sup> | NA | NA | NA | NA | NA | NA | NA |
| Vitule et al<sup>82</sup> | Present | - | - | - | - | - | NA |
| Waledziak et al<sup>83</sup> | - | - | - | - | - | - | NA |
| Xu et al<sup>84</sup> | Present x2 | - | - | Elevated ALT only x1 | Elevated x1 | - | Positive Murphy's sign x1, RUQ tenderness x1 |
| Yang et al<sup>85</sup> | Present x2 | - | - | ALT<br>elevated x2<br>and AST<br>elevated x3 | - | Elevated x2 | NA |
| Yoh et al<sup>86</sup> | - | - | - | - | - | - | NA |
| Yoshida et al<sup>87</sup> | - | - | - | - | Elevated | - | NA |
| Zheng et al<sup>88</sup> | - | - | - | - | - | - | NA |
| Zimpfer et al<sup>89</sup> | - | - | - | - | - | - | NA |

**Table 5:**
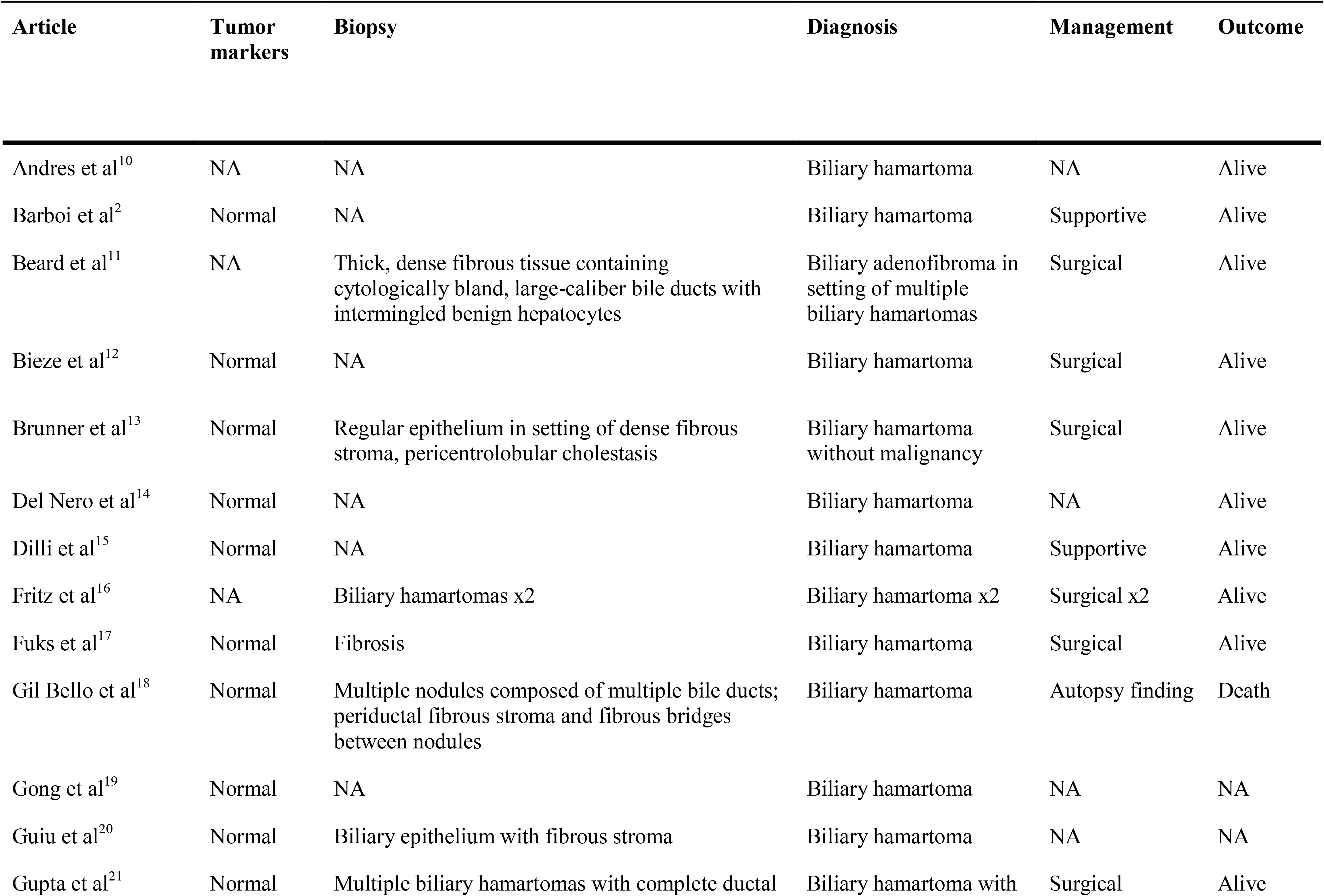
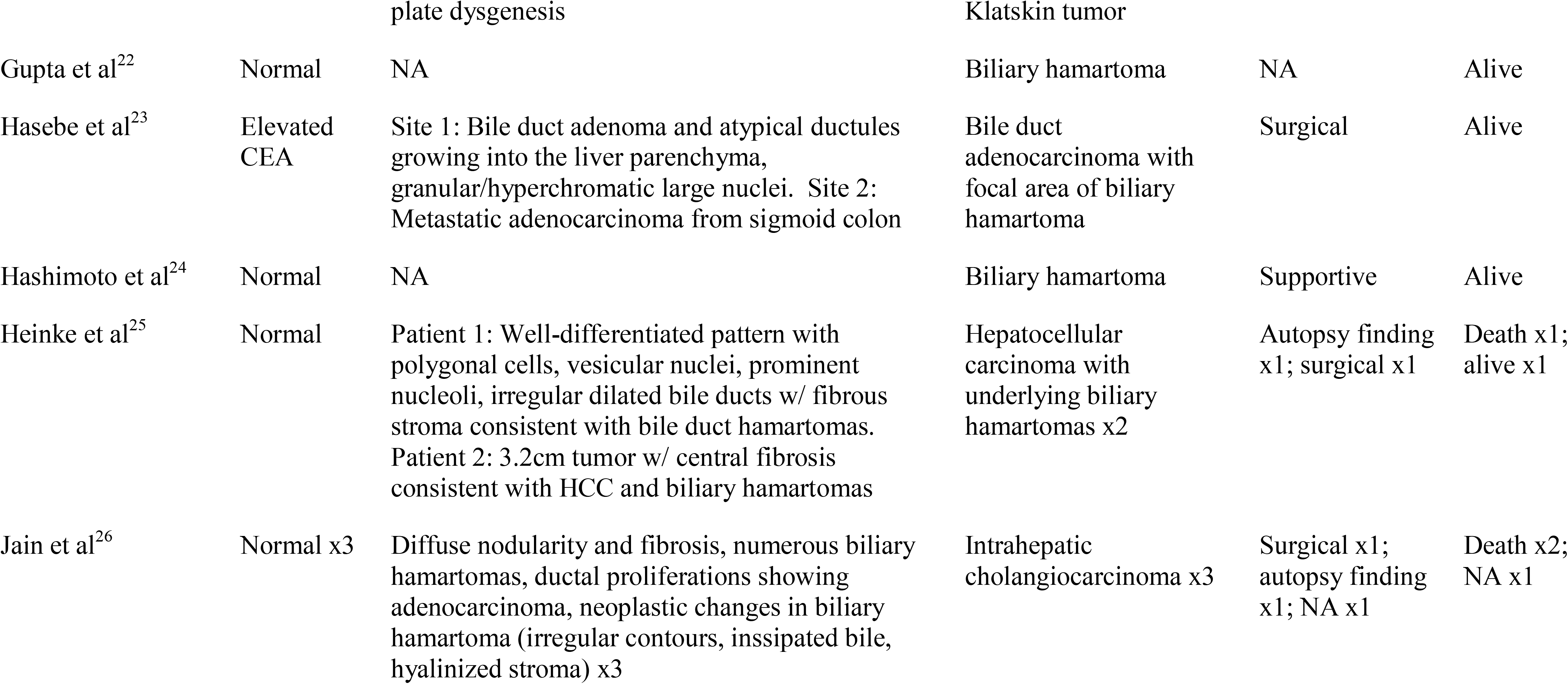
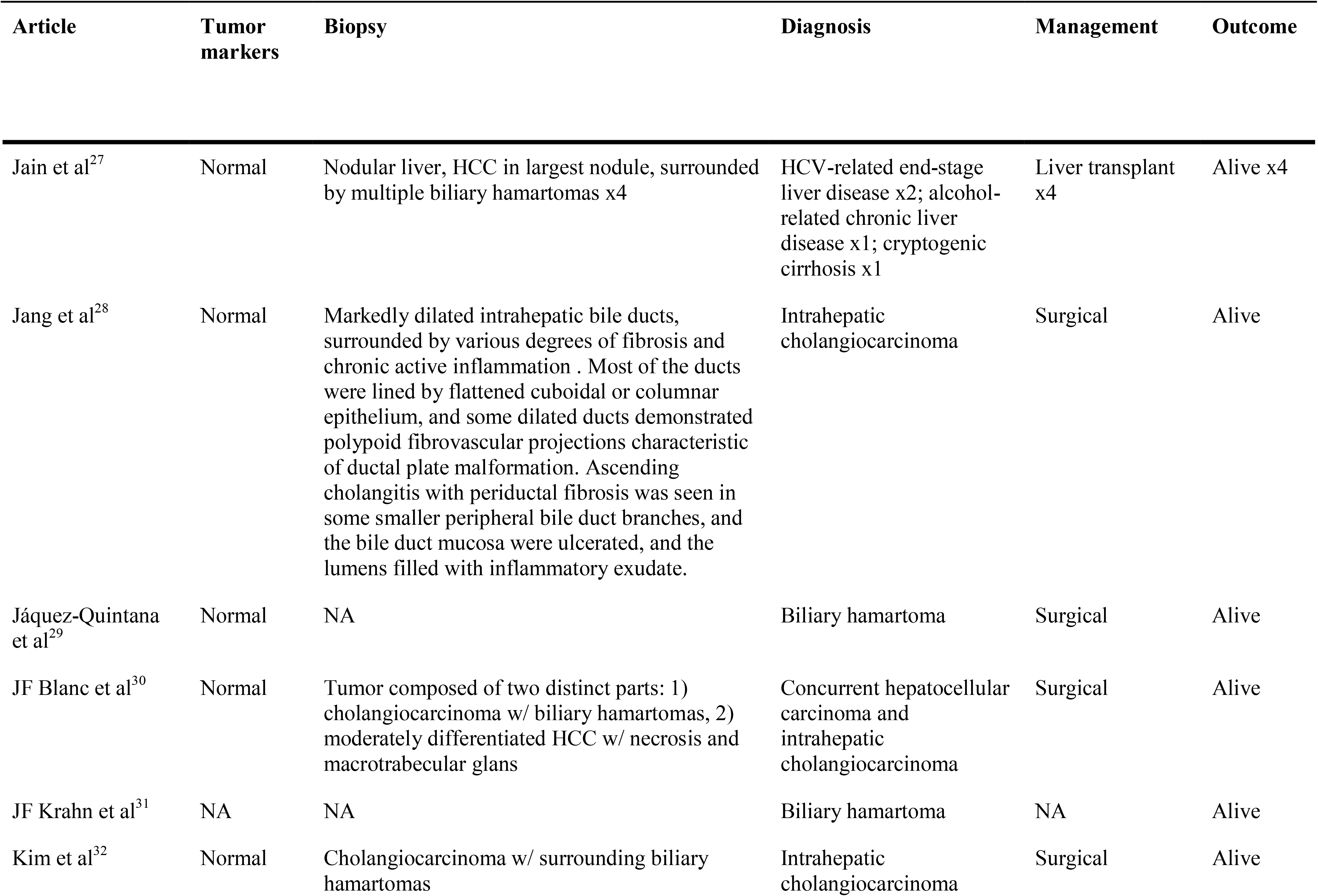
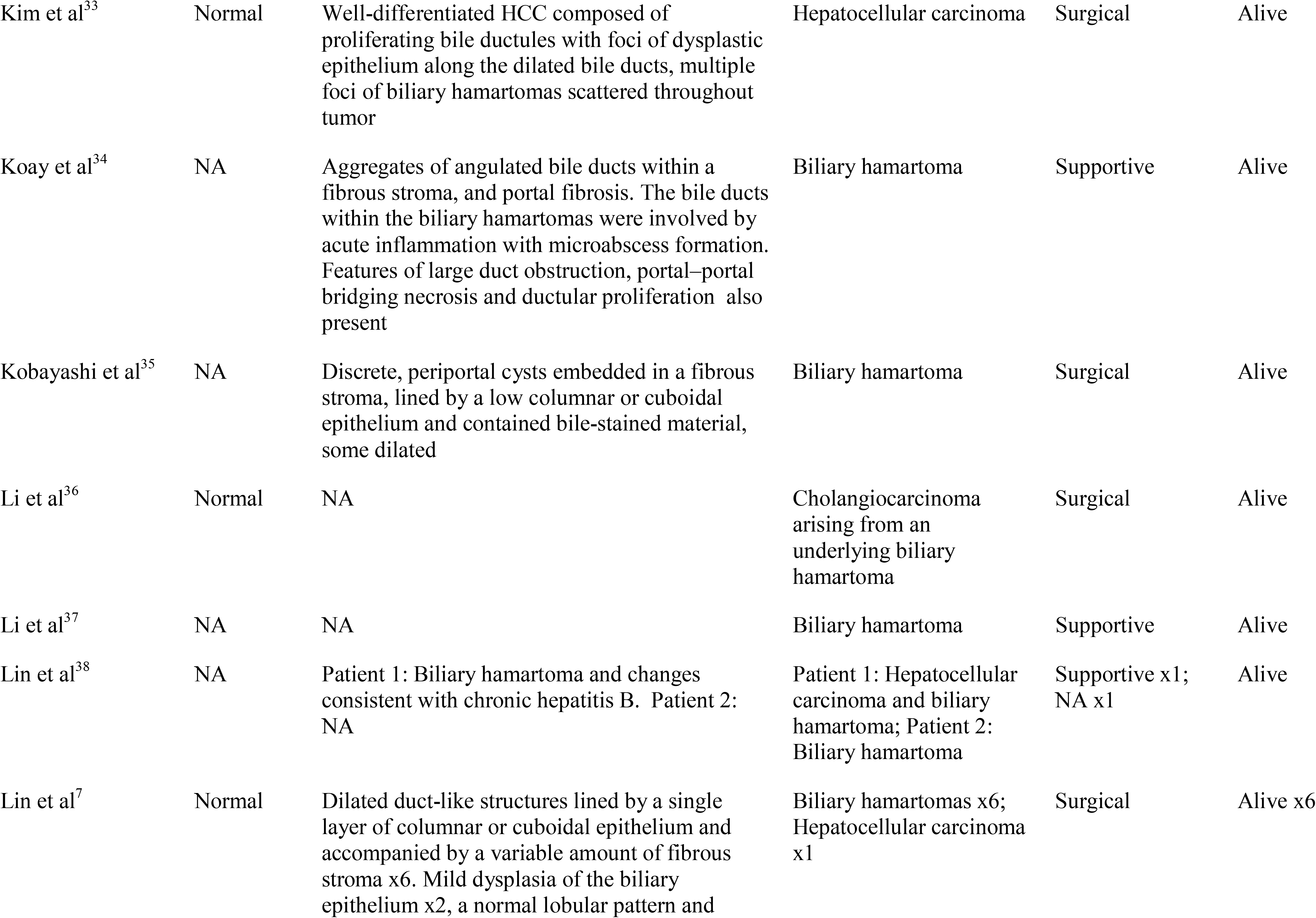
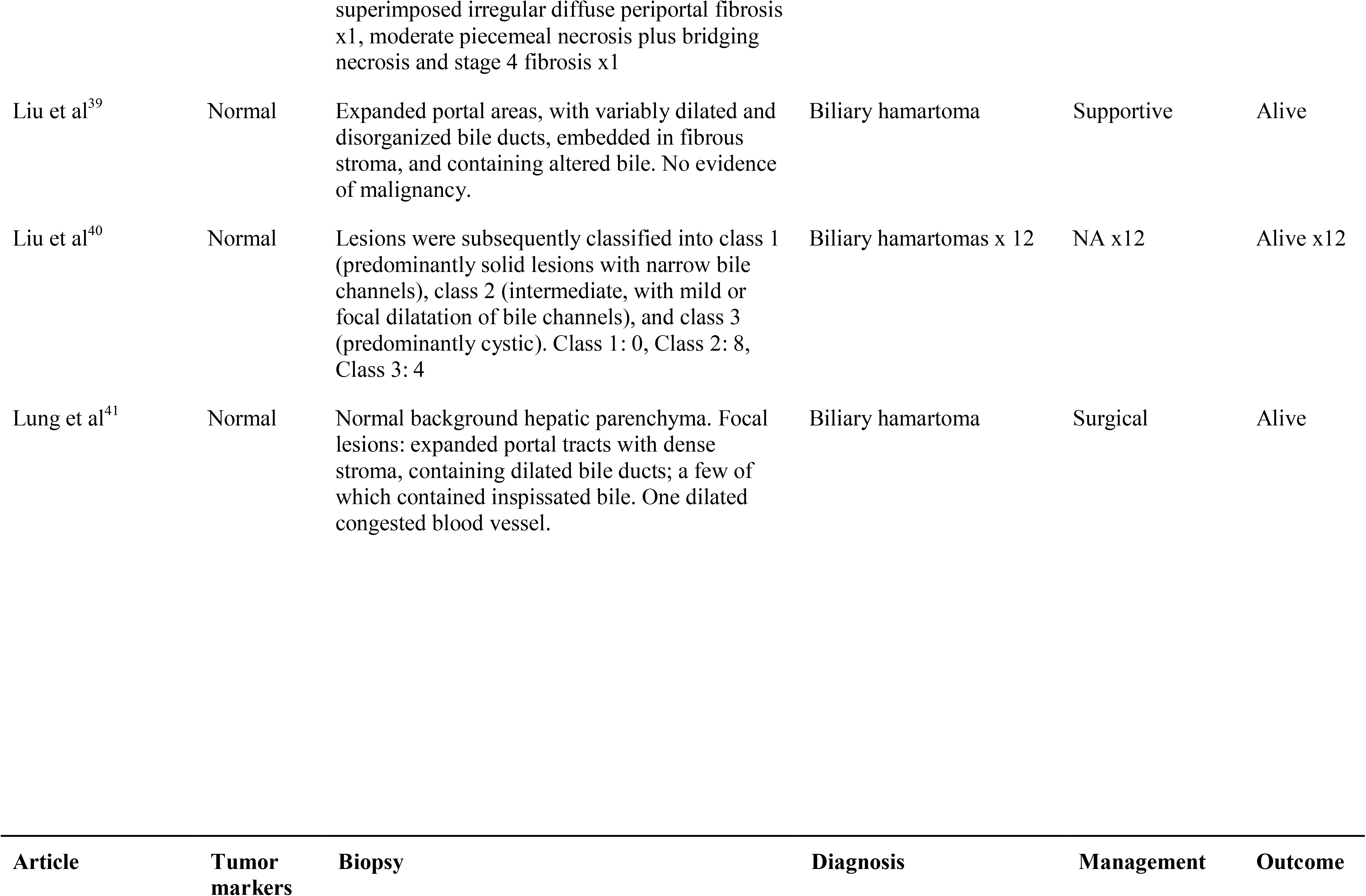
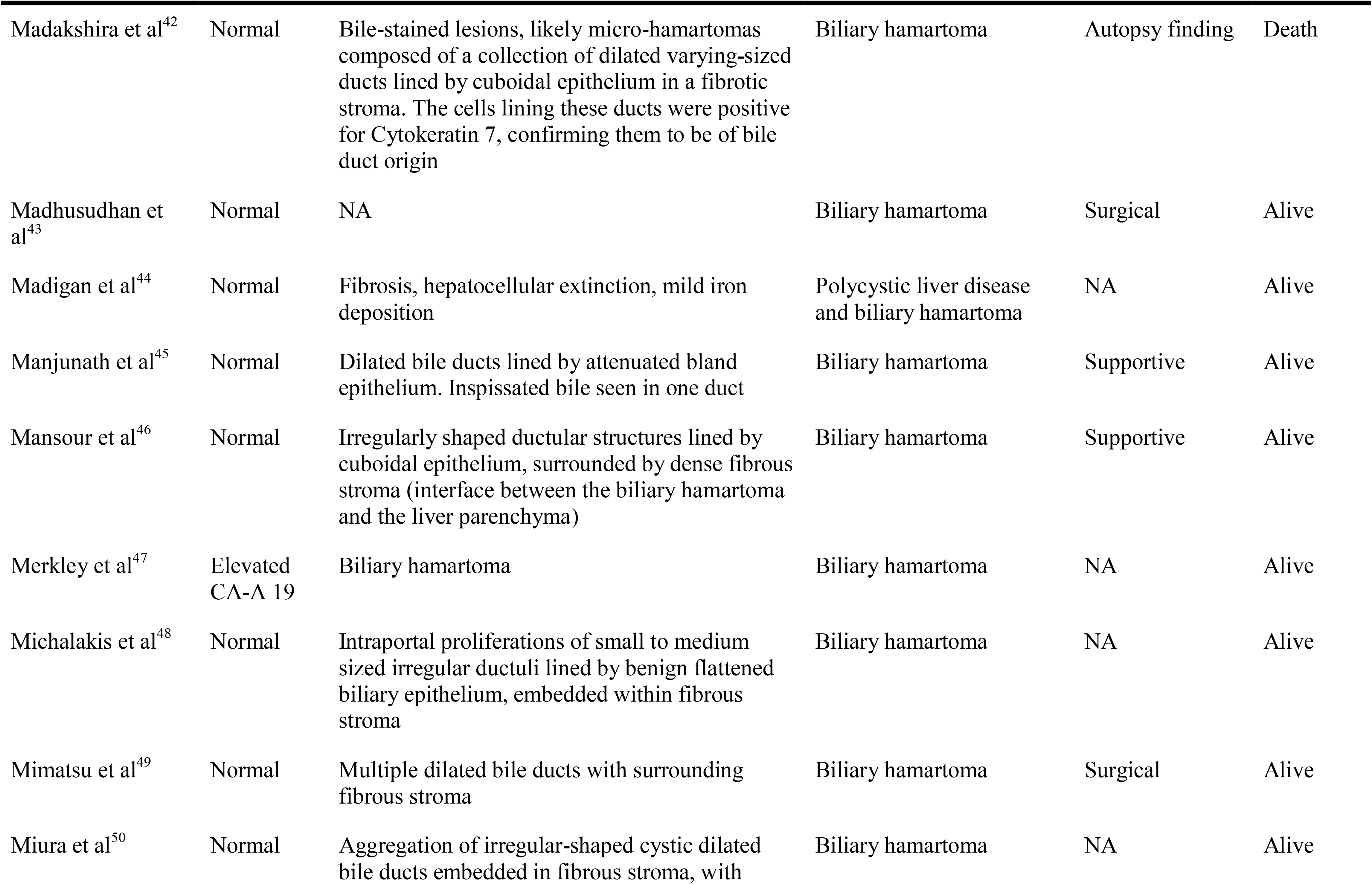
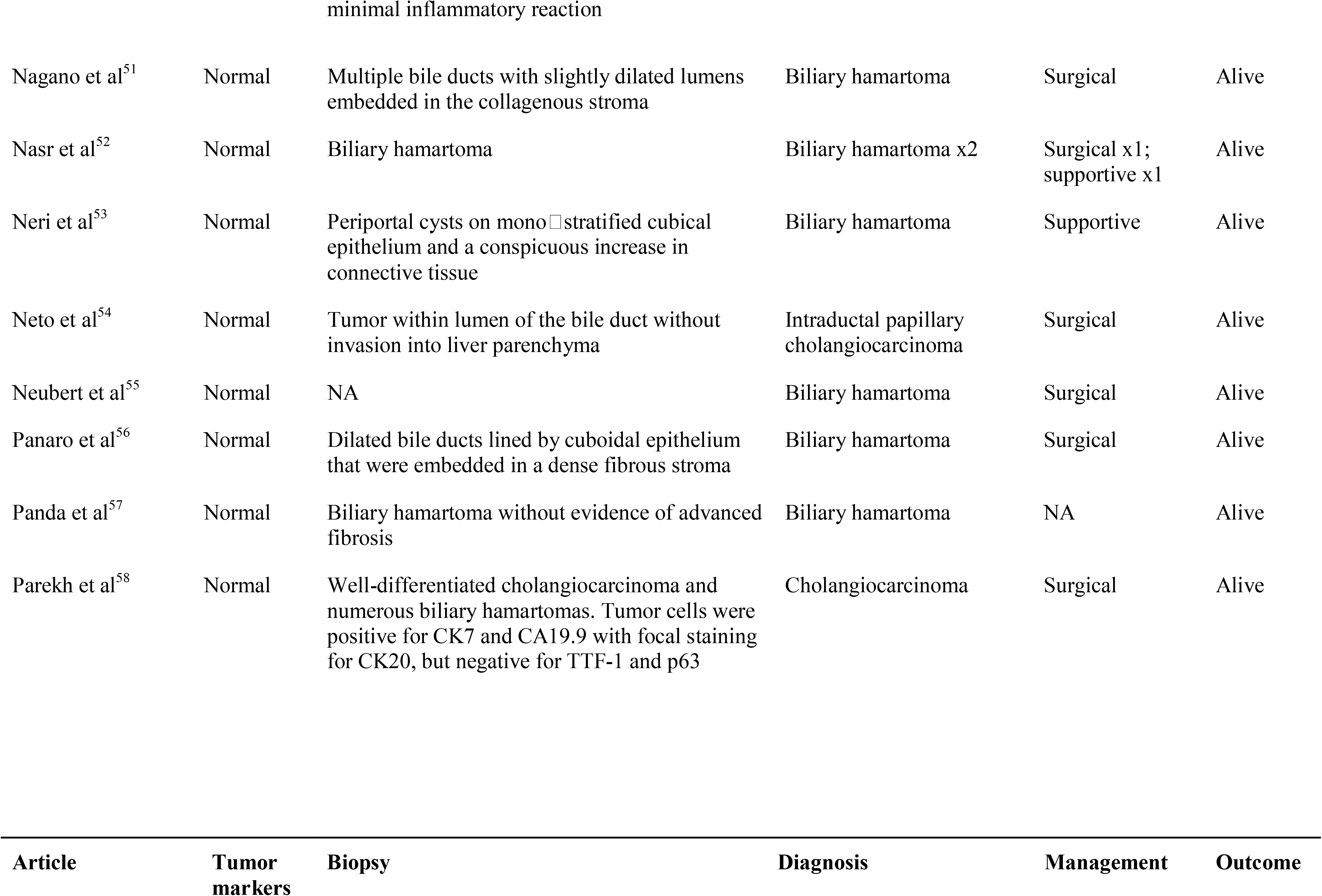
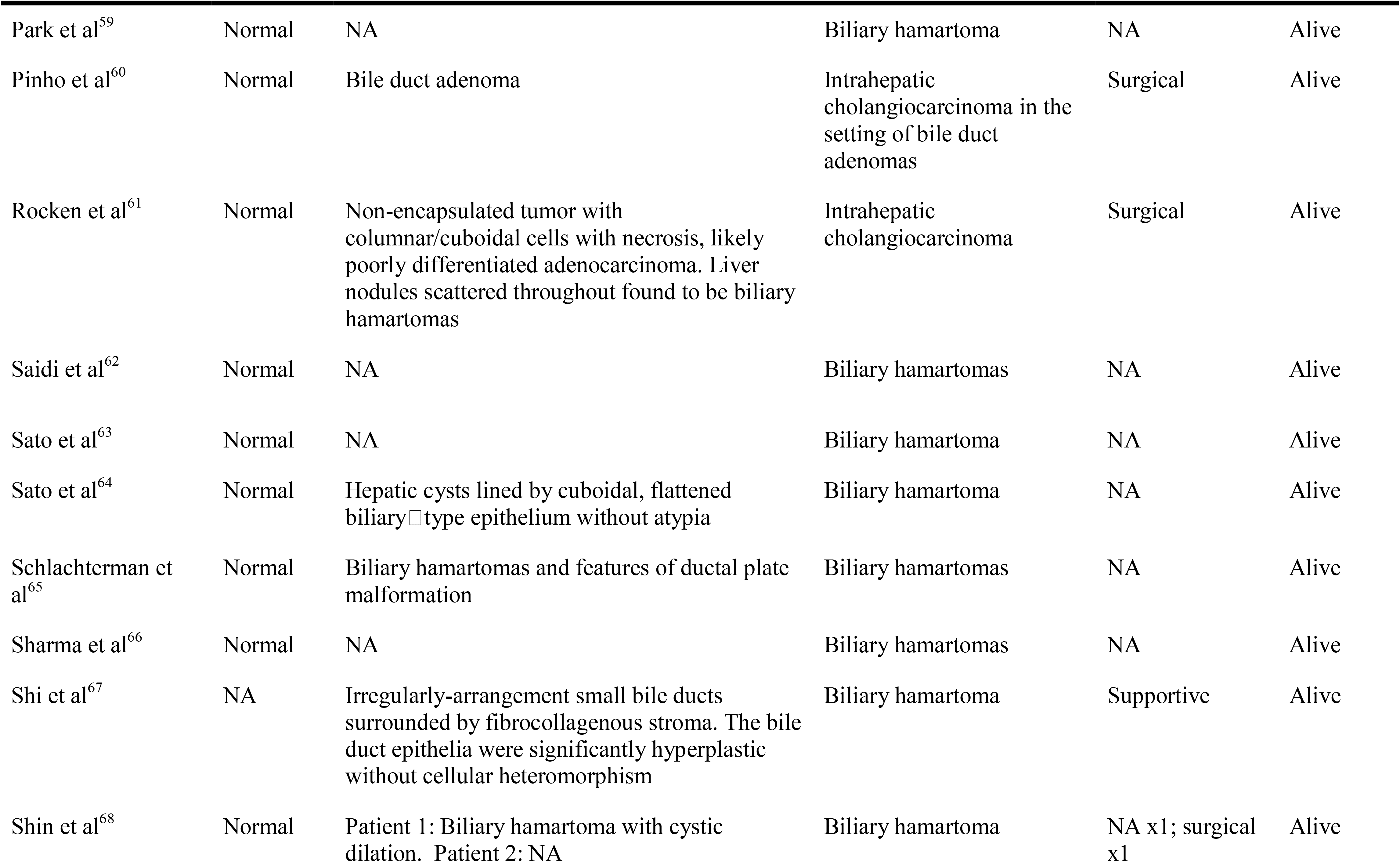
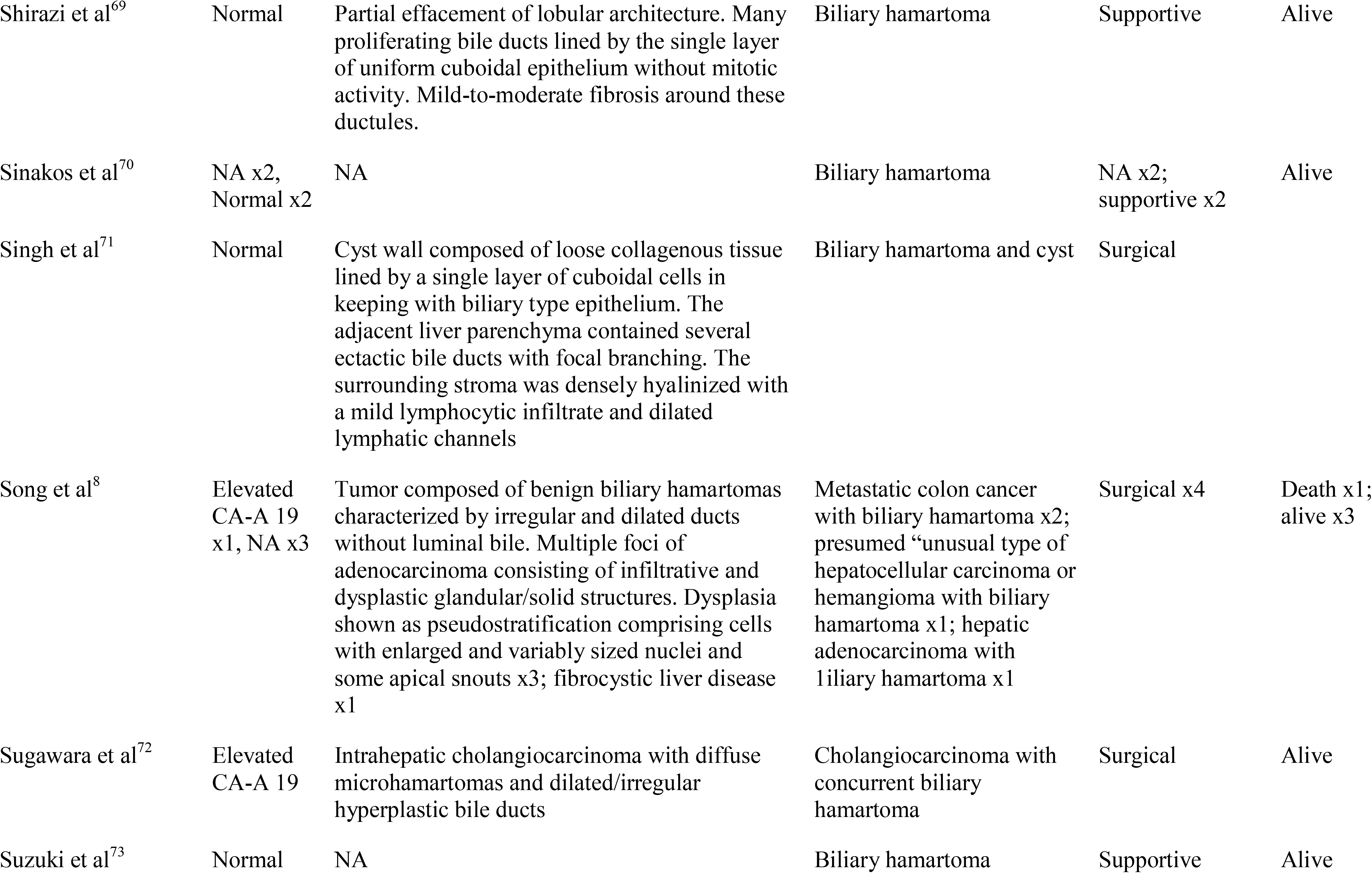
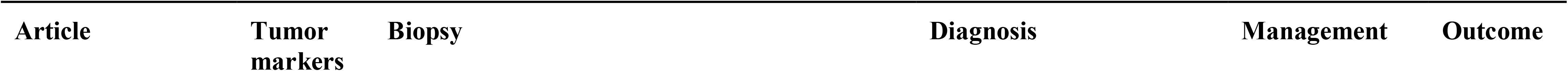
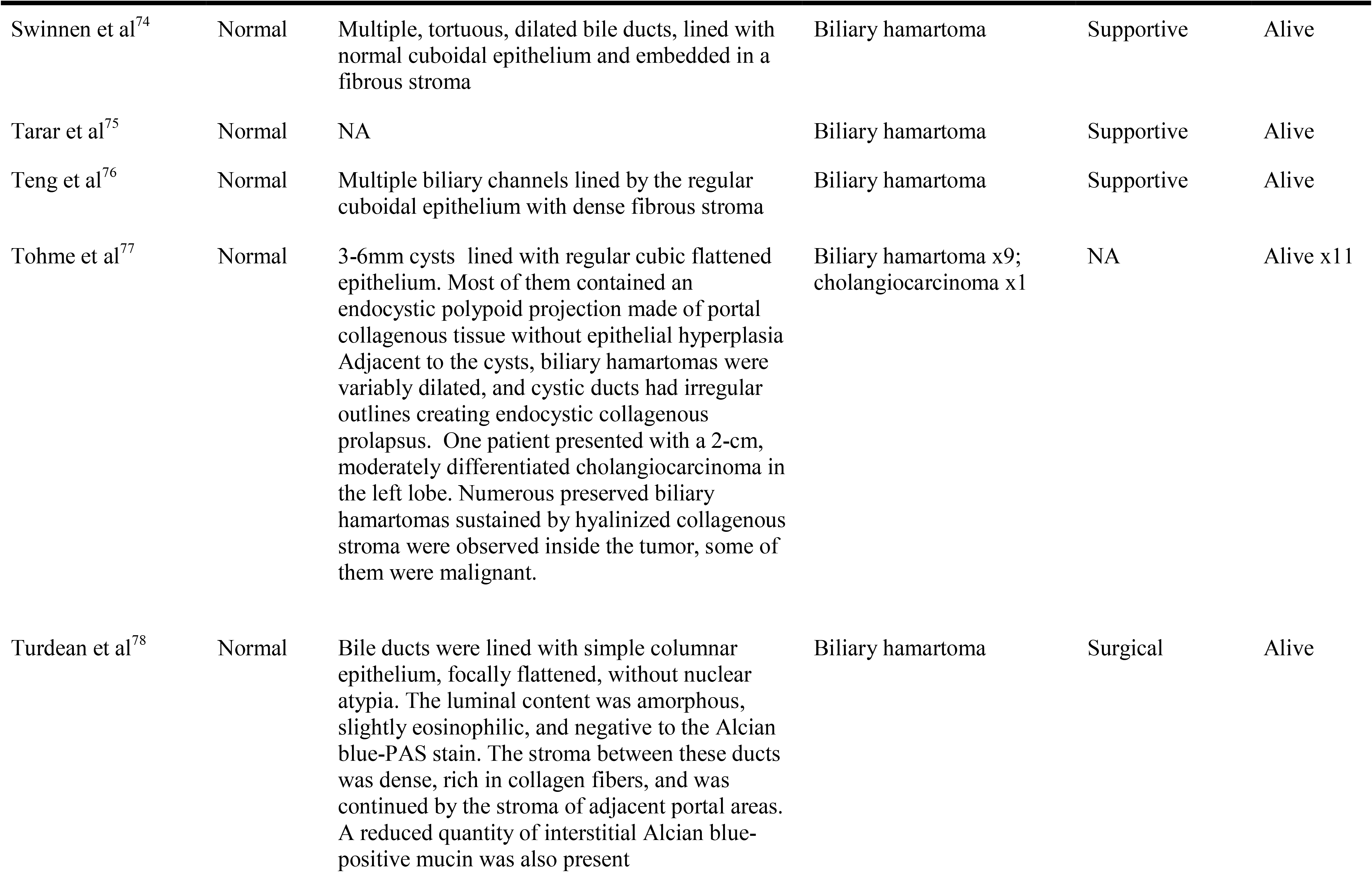
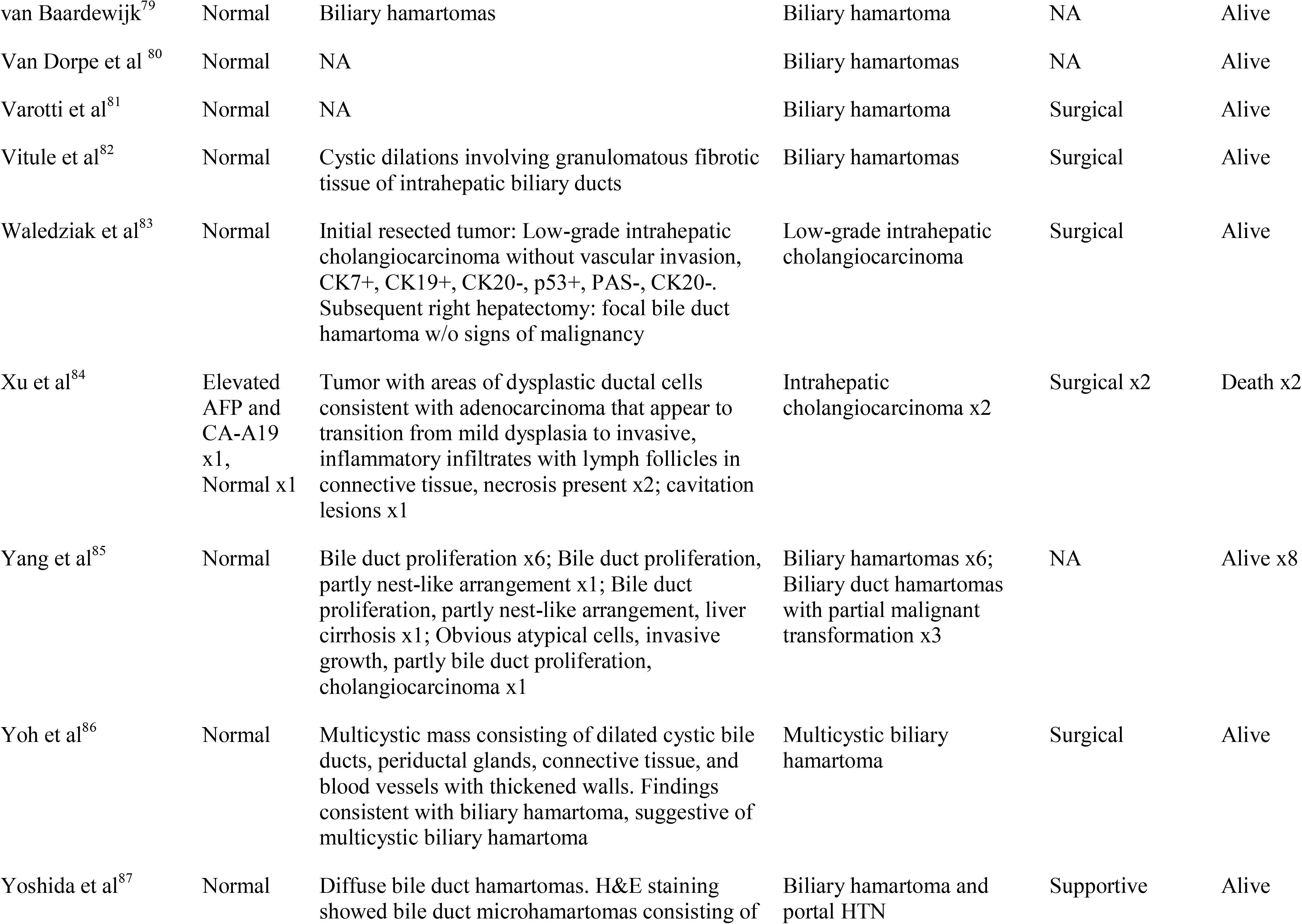
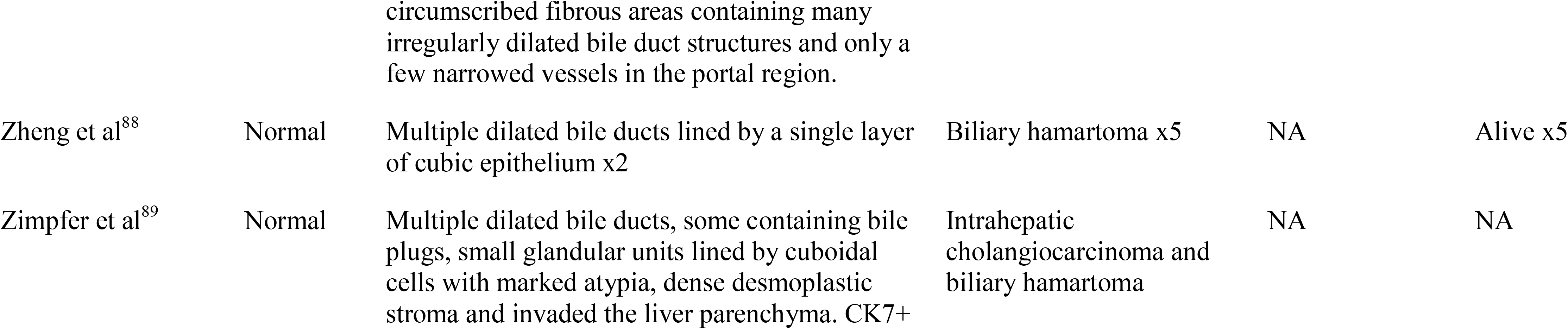
Pathological findings of cases reviewed.

### Patient Demographics

Patient ages ranged from 17 to 88 years at time of diagnosis, and the mean age of all patients in this analysis was 55±13 years. A slight male predominance was noted, with males comprising 63.3% (88 vs 51) of all cases reported.

### Comorbidities

Of 139 published cases, 48.9% had an underlying comorbidity reported. The most frequently reported comorbidity was cancer, which was present in 12.9% of patients. Among those cases, gastrointestinal cancers (e.g., esophageal cancers, gastric cancer, common bile duct cancer and colorectal cancer) were the most common, accounting for 12 of 18, or 66.7%, of underlying malignancies in this analysis. Comorbid malignancies of the breast, prostate, lung and thyroid were also reported.

Viral hepatitis was reported in 8.6% of patients in this analysis, with hepatitis B accounting for 7 of 12 cases (58.3%), and hepatitis C accounting for 5 of 12 of cases (41.7%). Additionally, 8.6% of patients were reported to have underlying hypertension, and 5.0% of patients had type 2 diabetes mellitus. Other reported comorbidities include tobacco use (4.3%), alcohol use (3.6%), hemochromatosis (1.4%) and tuberculosis (1.4%).

### Clinical Presentation

Only 36% of patients in this analysis were symptomatic at presentation, suggesting that majority (nearly 2/3) patients are asymptomatic at the time of presentation. Among patients who were symptomatic, abdominal pain was the most common symptom reported by 22.3% (31 of 139) of all patients and ranged from vague abdominal pain and discomfort to localized epigastric or right hypochondrial pain. Other commonly reported symptoms included: fever (18.0%), weight loss (14.0%), and jaundice (14.0%). Less frequently reported clinical features include abdominal distension and upper and lower gastrointestinal bleeding. The time from development of symptoms to presentation was variably reported among studies and ranged from one day to ten years.

Although clinical exam was also variably reported among studies, a positive Murphy’s sign and right hypochondrial tenderness to palpation were common among patients for whom a physical exam was reported (9 out of 26, or 34.6%), especially among patients who presented with abdominal pain. Hepatomegaly and splenomegaly were reported in a relatively small portion of patients with documented physical exams (19.2% and 13.6%, respectively).

### Laboratory Findings

Laboratory data, mainly alanine or aspartate transaminases (ALT/AST), was reported for 66 of 139 patients in this analysis. Of those 66 patients, 33.3% had some degree of elevated transaminase levels. Laboratory data for gamma-glutamyl transferase (GGT) and alkaline phosphatase levels (ALP) were reported for 71 patients and 42.2% of the patients (30 of 71) were reported to have elevated gamma-glutamyl transferase (GGT) or alkaline phosphatase levels. Laboratory data for bilirubin was reported for 60 patients in the analysis. Of these 60 patients, 15 patients (25.0%) had hyperbilirubinemia. Other reported laboratory abnormalities include hypoalbuminemia, elevated C-reactive protein (CRP) and hematologic abnormalities such as anemia, leukocytosis, leukopenia and thrombocytopenia.

Tumor marker data were reported for 42 of 139 patients. The vast majority (85.7%) did not have any tumor marker elevations. Four patients (9.5%) were found to have elevated levels of carbohydrate antigen-19 (CA-19), a tumor marker associated with gastrointestinal malignancies, especially pancreatic malignancies.^88, 90^ One patient (2.4%) was found to have elevated levels of alpha fetoprotein (AFP), a tumor maker associated with liver and testicular cancers, ^90, 91^ and one patient (2.4%) was found to have elevations in carcinoembryonic antigen (CEA), which is a tumor marker associated with malignancies in multiple organ systems, including the gastrointestinal system. ^90^

### Imaging

122 of 139 patients in this analysis underwent some form of abdominal imaging whereas the rest of the patients were diagnosed by biopsy results. 60 (43.2%) underwent ultrasound imaging, 69 (49.6%) underwent computed tomography (CT) scans, and 81 (58.2%) underwent magnetic resonance imaging (MRI) scans. Radiographic abnormalities were detected by all imaging modalities. The suspected lesions expected to be benign biliary duct hamartomas were hypoechogenic in 50.0% (30/60) of ultrasounds, hyperechogenic in 21.7% (13/60) whereas mixed echogenicity was seen in 28.3% (17/60). Similarly, for the same suspected lesions, CT radiography revealed hypointensity and hyperintensity in 81.2% (56/69) and 13.0% (9/69) of patients, respectively, whereas, 5.8% of the patients (4/69) had mixed intensity signals from the lesions. 40 CT scans utilized contrast, and of those, contrast-enhancement was present in 32.5% of images. Studies in this analysis reported both T1-weighted and T2-weighted MRI scans. Of the 81 T1- weighted scans performed for the suspected lesions, 96.3% reported hypointensity and 3.7% reported hyperintensity. In comparison, 6.2% of the 80 T2-weighted scans, for the benign lesions, reported hypointensity and 93.8% revealed hyperintensity. 41 MRI scans utilized contrast, and of those, 41.5% reported contrast-enhancement. Several patients also underwent magnetic resonance cholangiopancreatography (MRCP), which revealed hepatic lesions or cysts with no connection to the biliary tree. However on imaging, it was noted that in 3 cases, ultrasound revealed dilated biliary ducts. Furthermore, it was noted that in one case, CT scan revealed dilated biliary ducts. Similarly, the same finding was noted by MRI in 4 cases.

### Lesion Characteristics

Size and location of the lesions of interest were reported on radiology for 44.6% and 48.2% of cases, respectively. Of lesions for which size data is available, 53.2% were found to be <1cm in size. 22 of 67 lesions (32.8%) were found to be uniformly distributed throughout the liver, whereas 21 (31.3%) were confined to a particular segment of the liver. More lesions were found to be present in the right lobe of the liver than the left lobe (16 vs 6), and a small portion (2, or 3.0% of those with location information) were located on the surface of the liver. When visualized intraoperatively, the lesions were often described as grey or white nodular lesions, with some reports also describing a cystic component.

### Diagnosis, Management & Outcome

Most cases in this analysis employed biopsies as part of the diagnostic workup. Of the 112 cases for which biopsy results are available, the pathologic diagnosis was benign bile duct hamartoma in 69.6% of cases. In 8.9% of cases, the pathologic diagnosis was bile duct hamartoma with partial malignant changes, and in 21.4% of cases, the biopsy specimen was found to be consistent with malignancy.

Considering the final diagnosis reported in each case, the proportion of patients diagnosed radiologically with a malignancy was comparable to the pathology results; 26.6% of patients were diagnosed with some form of malignancy, whereas 73.3% were found to have a benign diagnosis.

Management strategies varied widely between cases. Of cases with a documented treatment approach, 59.0% proceeded with a form surgical management, most commonly partial hepatectomy. Other patients underwent cholecystectomies or liver transplants. In contrast, 36.1% of patients received supportive care. Among the patients who had received surgical intervention, 24.4% of the patients had been diagnosed with malignancies, commonly cholangiocarcinoma (14/20; 70.0%). A small portion of cases (4.8%) involved lesions found on autopsy, of which 2 patients had been diagnosed with malignancy (2/7; 28.6%).

Overall, outcomes were favorable for cases in this analysis. 116 cases reported patient outcomes; 107 cases (92.2%) reported that the patients were alive at time of publication, and 9 cases (7.8%), including those documenting lesions found on autopsy, reported that the patients were deceased at the time of publication. Outcomes were not documented for the remaining 23 cases.

## DISCUSSION

Biliary duct hamartomas are benign lesions lined by bile duct epithelium and often dilated with collections of bile in their lumen. Biliary duct hamartomas are commonly, hypoattentuating, non-enhancing lesions (Figure 2). These lesions vary in stromal characteristics and the presence of cystic dilatations of intrahepatic bile ducts covered by single layer of cuboidal cells with intervening fibrous stroma (Figure 3). Although rare, biliary duct hamartomas pose a diagnostic challenge because of their overlapping features with malignant masses.^3^ Therefore, our review aimed at describing common clinical characteristics, lab evaluation and imaging findings that could help in detecting biliary hamartomas when confronted with atypical hepatic lesions. In our systematic review, 82 studies were included based on the presence of biliary duct hamartomas. These 82 studies had 139 patients with biliary duct hamartomas presenting either as a symptomatic lesion, or as an incidental finding either on imaging or intraoperatively; or as a malignant transformation.

**Figure 2:**
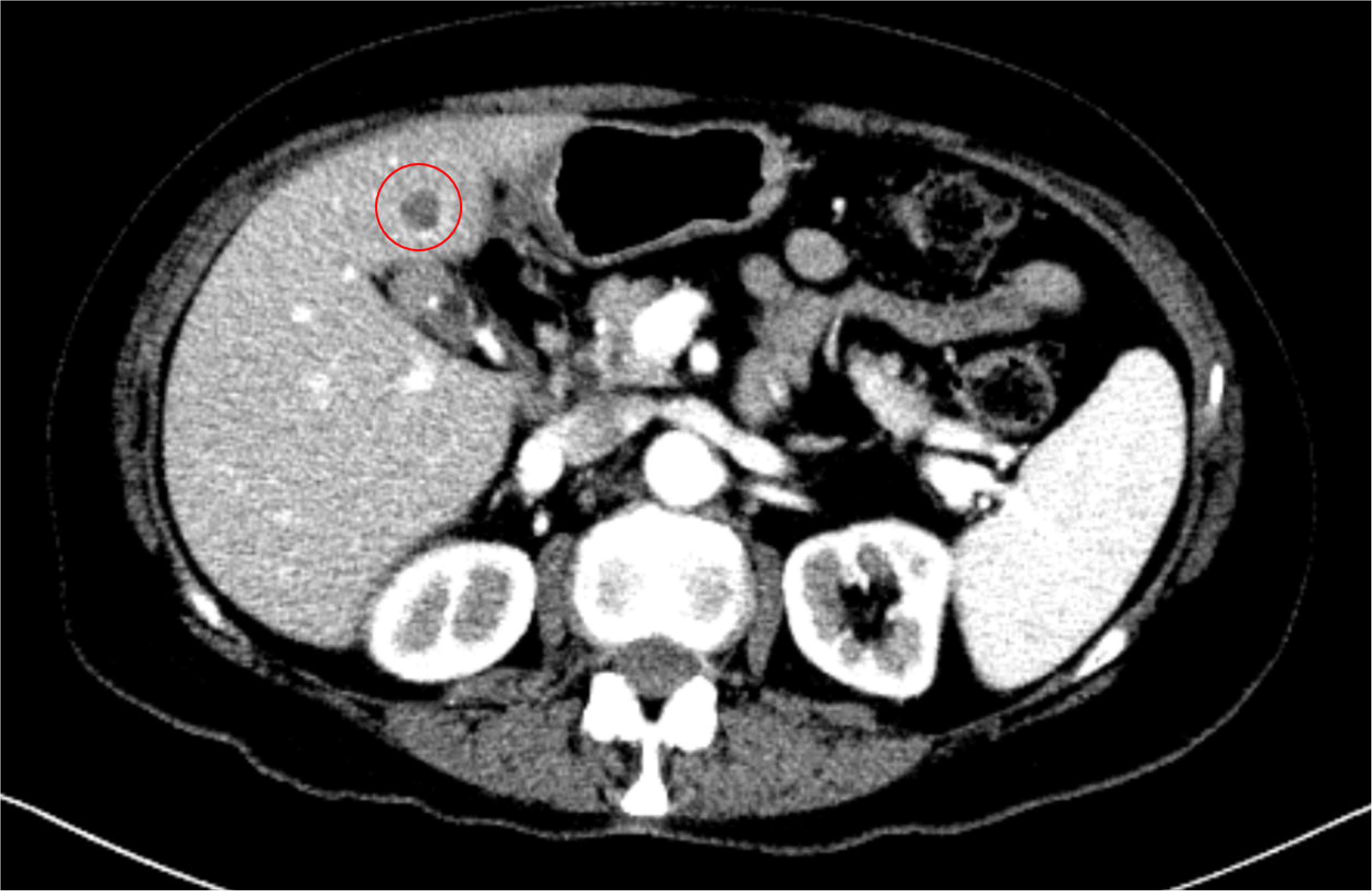
CT Abdomen Pelvis with contrast showing 1.1 cm hypodense lesion within segment IVb.

**Figure 3:**
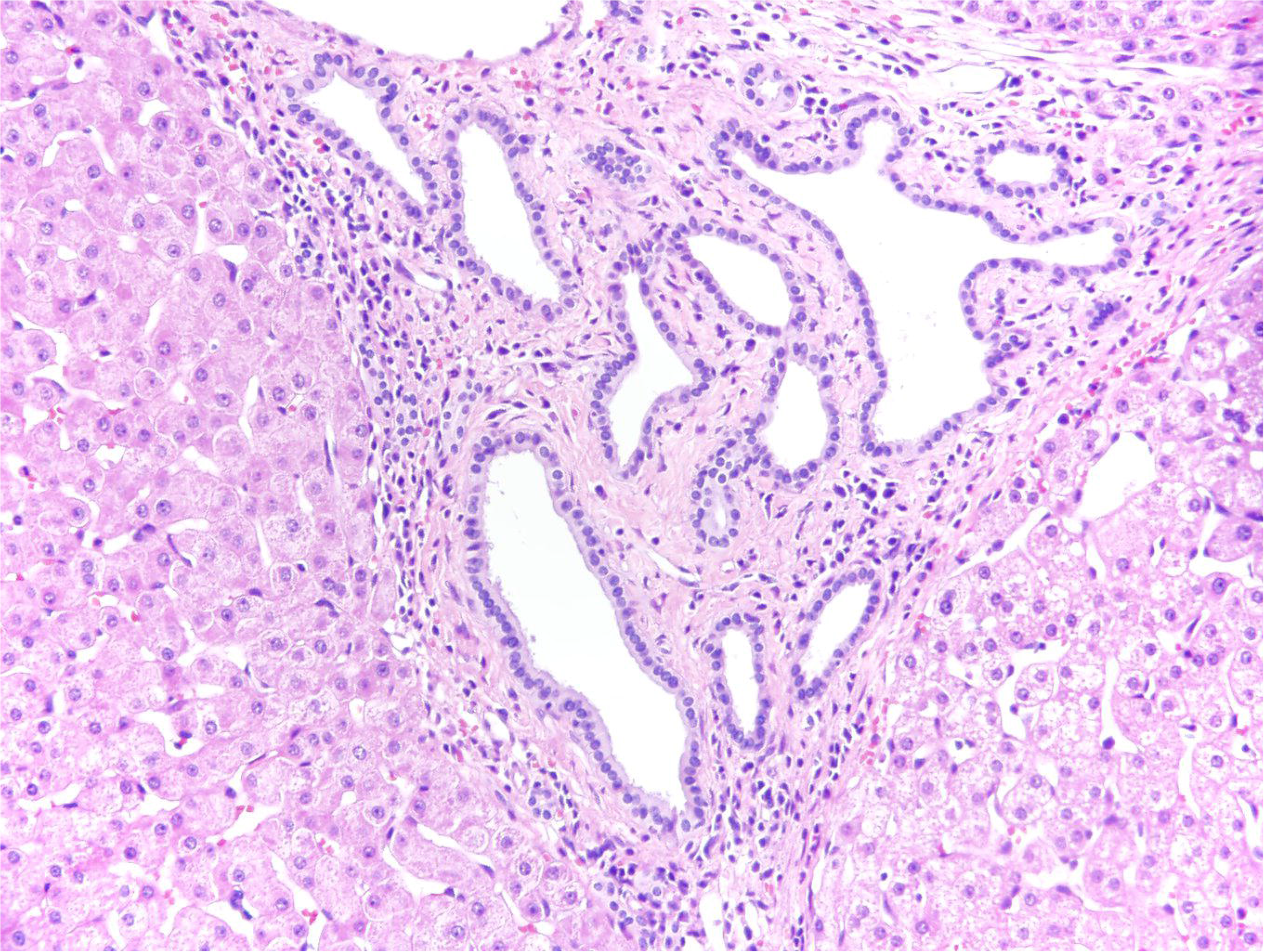
Liver parenchyma showing a well-demarcated lesion with small to medium sized, irregularly shaped dilated glands lined by bland cuboidal epithelium with intervening fibrous stroma and surrounding inflammatory cells consistent with biliary hamartomas (H&E, x20)

The patients’ ages ranged from 17 to 88 years, and the mean age of the patients was 55 ±13 years. Male predominance was noted (88 male patients vs. 51 female patients). The age range in our review was comparatively greater when compared to another study that mentioned biliary duct hamartomas as a diagnosis for patients more than 35 years of age.^7^ Furthermore, males were more likely to have such a mass as compared to females. This finding also differed from a study that determined female predominance for the condition.^7^

Comorbid conditions were present in approximately half of the patients with the commonest being malignancy (12.9%), particularly gastrointestinal malignancies followed by chronic viral hepatitis B and C (8.6%). This finding corroborates previous suggestions that biliary hamartomas are not necessarily congenital in nature but can also be acquired because of viral inflammation secondary to hepatitis B and hepatitis C.^3^ It is, however, important to note, that this increased frequency could also be observed due to selection bias, as patients with chronic liver disease are more likely to undergo abdominal imaging.

The majority of the patients were asymptomatic (64.0%); a finding that has been reported by multiple studies.^1, 88^ The most common symptom reported by the patients was abdominal pain followed by fever, weight loss, and jaundice. However, 41 symptomatic patients (82.0%) were diagnosed with biliary duct hamartomas whereas 9 symptomatic patients (18.0%) had been diagnosed with malignancies (cholangiocarcinoma, hepatocellular carcinoma and metastatic cancer). These symptoms have also been noted in other studies.^90, 91^

In our review, about one-third of 66 patients had elevated transaminases. Additionally, about one-fourth had hyperbilirubinemia. These elevations can suggest two possibilities; derangement in function test could occur as a result of obstruction of biliary network by a large lesion or transient damage to the lesion might manifest as the derangement.^92^ Tumor markers were reported to be normal for a majority of the 42/139 patients whose data was available (85.7%). This was similar to previous studies.^3–6^ However, a few exceptions were also found where CA-19, AFP, and CEA were elevated. This is most likely not specific to biliary hamartomas as elevated tumor markers are associated with many malignancies and might also be elevated in cases of underlying liver cirrhosis, portal hypertension, and jaundice. ^85^

Many common features were noted on ultrasonography. In most of the cases, the lesions were hypoechogenic (80%) and less than 1 cm in size (53.2%). This is an interesting finding, because a study noted that smaller lesions tend to be hyperechogenic in nature because of acoustic reflection from closely apposed walls.^40^ Therefore, our findings strengthen the prior idea that ultrasonography features were mostly non-specific for biliary duct hamartoma.^93, 94^ Lesions were mostly hypointense on CT-scan. Regarding contrast, few studies report that these lesions become clearer, instead of enhancing as found in our review (32.5%), with contras,; a confusing feature that might lead to a diagnosis of malignancy.^95, 96^ On MRI, most of the lesions produced hypointense and hyperintense signals on T1 and T2-weighted images respectively that has been mentioned in previous studies.^95, 96^ Interestingly, contrast enhancement was noted in 41.5% of the lesions, which can be attributed to compressed liver parenchyma surrounding the masses.^77^

Biliary duct hamartomas were usually found distributed randomly on the surface of the liver (32.8%) but were also confined to a segment (31.5%). The right lobe was more involved than the left lobe. The reason for the difference in the involvement of lobes is not known. However, diffuse distribution of nodules on the surface of the liver was considered a feature suggestive of a benign form of biliary duct hamartomas.^95^

The best diagnostic tool was, indeed, a biopsy, performed in 112 patients. The majority of the patients had a benign pathological diagnosis of biliary duct hamartomas (73.3%) but 59.9% still underwent some form of surgical procedure for management.

As mentioned in many studies, the outcomes were generally favorable with many patients surviving long-term unless they had other conditions that could potentially aggravate their condition in the long run.^1, 92^ This is true of hamartomas across organ systems— hamartomas of the lungs, small bowel and pancreas have been reported to have generally favorable outcomes.^96–98^ Apart from the negligible risk of malignant transformation, biliary hamartomas are a benign condition with no known long-term adverse outcomes. Consequently, no intervention is required in an asymptomatic patient.^1^

A major strength of this review is the high number of patients, which allowed for a thorough analysis of the clinical, diagnostic, and prognostic features of biliary duct hamartomas. One potential limitation is publication bias—cases that involve significant or unusual symptoms or that are associated with malignant transformation are more likely to be reported in the literature than clinically inconsequential cases.

## CONCLUSION

In summary, this systematic review of 82 studies and 139 patients represents the largest cohort of patients with biliary duct hamartomas evaluated to date. Biliary duct hamartomas can be associated with mild symptoms and lab derangements. Despite being detectable on imaging, findings are non-specific and might necessitate the use of biopsy. Currently, there are no published guidelines regarding the management of biliary duct hamartomas, and many patients have undergone surgical management despite the clinically benign nature of these malformations. Further inquiry is necessary to define the optimal management of this uncommon condition.

## Data Availability

Data will be made available on special request by the corresponding author.

